# Serum anti-NMDA-receptor antibodies and cognitive function after ischemic stroke (PROSCIS-B)

**DOI:** 10.1101/2020.04.15.20066324

**Authors:** Pia S. Sperber, Pimrapat Gebert, Leonie H.A. Broersen, Shufan Huo, Sophie K. Piper, Harald Prüss, Peter U. Heuschmann, Matthias Endres, Thomas G. Liman, Bob Siegerink

**Affiliations:** Charité – Universitätsmedizin Berlin, corporate member of Freie Universität Berlin, Humboldt-Universität zu Berlin, and Berlin Institute of Health, Center for Stroke Research Berlin (CSB), Berlin, Germany; German Centre for Cardiovascular Research DZHK, partner site Berlin, Germany; Charité – Universitätsmedizin Berlin, corporate member of Freie Universität Berlin, Humboldt-Universität zu Berlin, and Berlin Institute of Health, Institute of Biometry and Clinical Epidemiology, Berlin, Germany; Berlin Institute of Health (BIH), Charité – Universitätsmedizin Berlin and Max Delbrück Center for Molecular Medicine in the Helmholtz Association, Berlin, Germany; Charité – Universitätsmedizin Berlin, corporate member of Freie Universität Berlin, Humboldt-Universität zu Berlin, and Berlin Institute of Health, Department of Neurology with Experimental Neurology, Berlin, Germany; German Center for Neurodegenerative Disease DZNE, partner site Berlin, Germany; University of Würzburg, Institute of Clinical Epidemiology and Biometry, Würzburg, Germany; University Hospital Würzburg, Clinical Trial Center Würzburg, Würzburg, Germany; Charité – Universitätsmedizin Berlin, corporate member of Freie Universität Berlin, Humboldt-Universität zu Berlin, and Berlin Institute of Health, Excellence Cluster Neurocure, Berlin, Germany

**Keywords:** stroke, ischemia, epidemiology, antibodies, cognitive dysfunction

## Abstract

**Objective:** We aimed to investigate whether serum anti-N-Methyl-D-Aspartate-receptor GluN1 antibodies (NMDAR1-abs) are associated with worse cognitive function over three years after first ischemic stroke.

**Methods:** Data were used from the PROSpective Cohort with Incident Stroke-Berlin (PROSCIS-B;NCT01363856). NMDAR1-abs (IgM/IgA/IgG) were measured with cell-based assays from serum obtained within seven days after first-ever stroke. Seropositivity was defined as titers ≥1:10, low titers as ≤1:100 and high titers as >1:100. We assessed cognitive status annually up to three years after stroke with the Telephone Interview for Cognitive Status – modified (TICS–m) and used crude and adjusted linear mixed models to estimate the impact of NMDAR1-abs exposure on TICS-m over time.

**Results:** Data on NMDAR1-abs (median day of sampling=4[IQR=2–5]) were available in 583 of 621 PROSCIS-B patients (39% female; median NIHSS=2[IQR=1–4]; median MMSE=28[IQR:26–30]; median mRS=2[IQR=1–3]) of whom 76(13%) were seropositive (IgM:n=48/IgA:n=43/IgG:n=2). TICS-m over time was not different in NMDAR1-abs seropositive compared to seronegative patients (β_Crude_=0.38[95%CI=-1.00 to 1.76]; adjusted β_Model3_=0.30[95%CI=-1.14 to 1.73]). In subgroups, TICS-m over time was not different in patients with low titers (β_Crude_=1.53[95%CI=-0.06 to 3.11]; adjusted β_Model3_=1.42[95%CI=-0.23 to 3.08]), however, in patients with high titers TICS-m was lower in the crude model (β=-2.54[95%CI=-4.99 to -0.08]), with a similar effect size after confounder adjustment (β_Model3_=-2.30[95%CI=-4.82 to 0.21]). All groups were compared to seronegative patients, respectively.

**Conclusion:** Overall NMDAR1-abs seropositivity was not associated with cognitive function over time after first-ever mild-to-moderate ischemic stroke. Our data suggest that high titers associate with impaired cognitive function after stroke, warranting larger studies.

## Introduction and Background

Cognitive impairment is frequent after stroke and up to one third of all stroke patients develop incident post stroke dementia.(1,2) Serum anti-NMDA (N-Methyl-D-Aspartate)-receptor antibodies (NMDAR1-abs), primarily of the IgA and IgM isotypes have been observed in about 10% of the apparently healthy population, however, other studies also showed associations between seropositivity and cognitive impairment.(3–5) We previously observed that NMDAR1-abs seropositivity with high titers (>1:100) was associated with unfavorable functional outcome at one year after stroke and overall seropositivity was associated with increased vascular recurrent risk or death within three years after ischemic stroke,(6) contrasting previous findings that indicated beneficial effects of serum NMDAR1-abs on infarct volumes in ischemic stroke.(7,8) The functionality of IgA and IgM antibodies has been studied previously and remains controversial, especially the clinical implication of these isotypes.(9,10) Physiological NMDAR function is crucial for mechanisms of synaptic plasticity and normal neuronal function.(11) We consider that serum NMDAR1-abs may enter brain parenchyma following blood brain barrier disruption as a consequence of stroke where they may lead to a downregulation of NMDA-receptors and hamper functional recovery of the damaged tissue.(10) Therefore, this study was motivated by the hypothesis that NMDAR1-abs have detrimental effects on cognitive outcome after stroke. We herewith aimed to quantify the impact of NMDAR1-abs on cognitive functioning up to three years in a large cohort of first-ever ischemic stroke patients.

## Material and Methods

### The PROSpective Cohort with Incident Stroke - Berlin (PROSCIS-B) study

The PROSCIS – B study (ClinicalTrials.gov identifier: NCT01363856) is a prospective observational hospital-based cohort study, which recruited patients at three tertiary university hospital stroke units of the Charité–Universitätsmedizin Berlin with first-ever stroke according to WHO criteria, to study stroke secondary risks.(12) Patients presenting with brain tumor, brain metastasis of a tumor of other origin, or patients participating in an intervention study, were excluded. Details on the study design have been described previously.(13,14) For a detailed baseline characterization, an extensive clinical and technical examination was performed within seven days after the acute event including blood sampling for laboratory measures. Patients were followed-up annually by telephone interviews or postal mail contact assessing i.a. cognitive function and functional outcome up to three years after the index event. For this investigation, only patients with mild-to-moderate ischemic stroke events (National Institutes of Health Stroke Scale [NIHSS] < 16) were included, as we counted very few cases with severe strokes (NIHSS > 15, n=6).

### Assessment of anti-NMDA-R antibodies

Serum blood samples were obtained from patients within 7 days after stroke and stored at −80°C until they were first-ever thawed for antibody measurements. NMDAR1-abs were measured with cell-based assays by the Euroimmun laboratory in Luebeck, Germany. In summary, HEK293 cells were transfected with GluN1 subunits of NMDA-receptors to bind antibodies of the IgM, IgA and IgG isotype from patient serum. Fluorescein isothiocyanate anti-human IgG was secondary administered to manually obtain staining with fluorescence microscopy. The assessors had no insight to patient data. Details on the procedure have been described elsewhere.(15,16) Titer levels started from a dilution of 1:10, which defined seropositivity in our study. For sub-groups, we defined titers of 1:10 to 1:100 as low titers and titers > 1:100 as high titers a priori, in line with previous analyses.(6)

### Outcome definitions

Cognitive function at baseline was measured with the Mini Mental State Examination (MMSE) and cognitive impairment at baseline was defined as MMSE <26.(17) For our main outcome of interest we used the validated German version of the Telephone Interview for Cognitive Status-modified (TICS-m) to annually assess cognitive status after stroke.(18) TICS-m is a screening instrument for cognitive impairment consisting of 20 questions, the points of which add up to a maximum of 50 points total. We provide all requested items for this instrument in the online supplement (Supplemental Methods I). The first TICS-m assessment was at one year and the last one about three years after stroke.

### Statistical methods

For our main analyses, we set up linear mixed models (LMM) to estimate the impact of NMDAR1-abs seropositivity on TICS-m over time in comparison to seronegative patients, who acted as the reference group for all analyses in this study. Time in years on a continuous scale and NMDAR1-abs exposure were used as fixed effects in the model equation, whereas individual identification (patient ID) was modelled as random effect. This random intercept mixed model allowed individuals within an exposure group to start from different basic cognitive levels without violating the overall group effect for seropositivity and respective subgroups in comparison to seronegativity on TICS-m scores over time. The estimated effect sizes (β) for our exposure groups indicate the additional increase or decrease of TICS-m points regarded over the three years. The full equation can be extracted from Supplemental Methods II (online only). We calculated 95% confidence intervals in addition to our effect estimate as a measure of precision from our sample.

Potential confounders were selected a priori according to their presumable impact on both, NMDAR1-abs serostatus and post-stroke cognitive functioning. We adjusted our first model (Model1) for age, sex and years of school education in 2 categories (≤ 10 years of school;>10 years of school), the second Model (Model2) was adjusted for a propensity score, which was built using the variables age, sex, years of school education, current smoking (yes/no), and a disease history assessment requesting history of 1) arterial hypertension (yes/no), 2) peripheral artery disease (yes/no), 3) atrial fibrillation (yes/no), 4) diabetes mellitus (yes/no), and 5) myocardial infarction (yes/no). For the binary exposure status (seropositive vs. seronegative) we built the propensity score with a logistic regression model and for the exposure in ordered subgroups, i.e. seronegative patients, patients with low titers and patients with high titers, we used an ordered logistic regression model.(19) In a third model (Model3) we adjusted for stroke severity (NIHSS in 2 categories: 5-15 vs. 0-4) and stroke etiology classified by the Trial of Org 10172 in Acute Stroke Treatment (TOAST)-criteria (1.large-artery atherosclerosis, vs. 2.cardioembolism /3.small vessel occlusion /4.other cause / 5.unknown etiology) in addition to the propensity score. This approach was to evaluate the robustness of our effect estimate after incorporating these attributes. The methodological approach of our sensitivity analyses in which we 1) explored the pattern of missing TICS-m outcome values, 2) applied multiple imputation by chained equations (MICE) on missing values and 3) estimated effects after excluding TICS-m outcomes of depressed patients are provided in Supplemental Methods III and Supplemental Methods IV.

Data preparation was done in IBM SPSS Statistics for Windows, version 24 (IBM Corp., Armonk, N.Y., USA). Data visualization was conducted in R i386 3.5.1, the R foundation, with the RStudio interface using the ggplot2 package. All statistical analyses were performed using Stata version 14.2 (Stata Corp., College Station, TX, USA).

### Ethics approval

All patients or their legal guardian gave written informed consent for study participation. PROSCIS-B was approved by the local ethics committee of the Charité – Universitätsmedizin Berlin and the study was conducted in concordance to ethical principles framed by the declaration of Helsinki.

### Data availability

The data that support the findings of this study are available from the qualified principal investigator of PROSCIS-B (T.G. Liman) upon reasonable request.

## Results

### Main results

PROSCIS-B recruited patients between March 2010 and February 2013 at three campuses of the Charité – Universitätsmedizin Berlin, of whom 621 presented with mild-to-moderate ischemic stroke and were thus eligible for analyses. Of those, serum samples were available in 583 patients and the median day of blood sampling from index stroke was 4 (IQR=3 to 5) in overall seropositive patients, 4 (IQR=3 to 6) in patients with low titers and 4 (IQR=3 to 5) in patients with high titers.(6) Details on patient inclusion and exclusion are presented in the flowchart in Figure 1, which further provides an overview of missing TICS-m observations at each year. At least one TICS-m score was available in 452 (451 of those 583 with antibody measurement) patients and 169 patients were missing all TICS-m follow-up assessments. Fifty-five patients died during the three-year follow up period and were therefore missing single or multiple TICS-m assessments.

**Figure 1:**
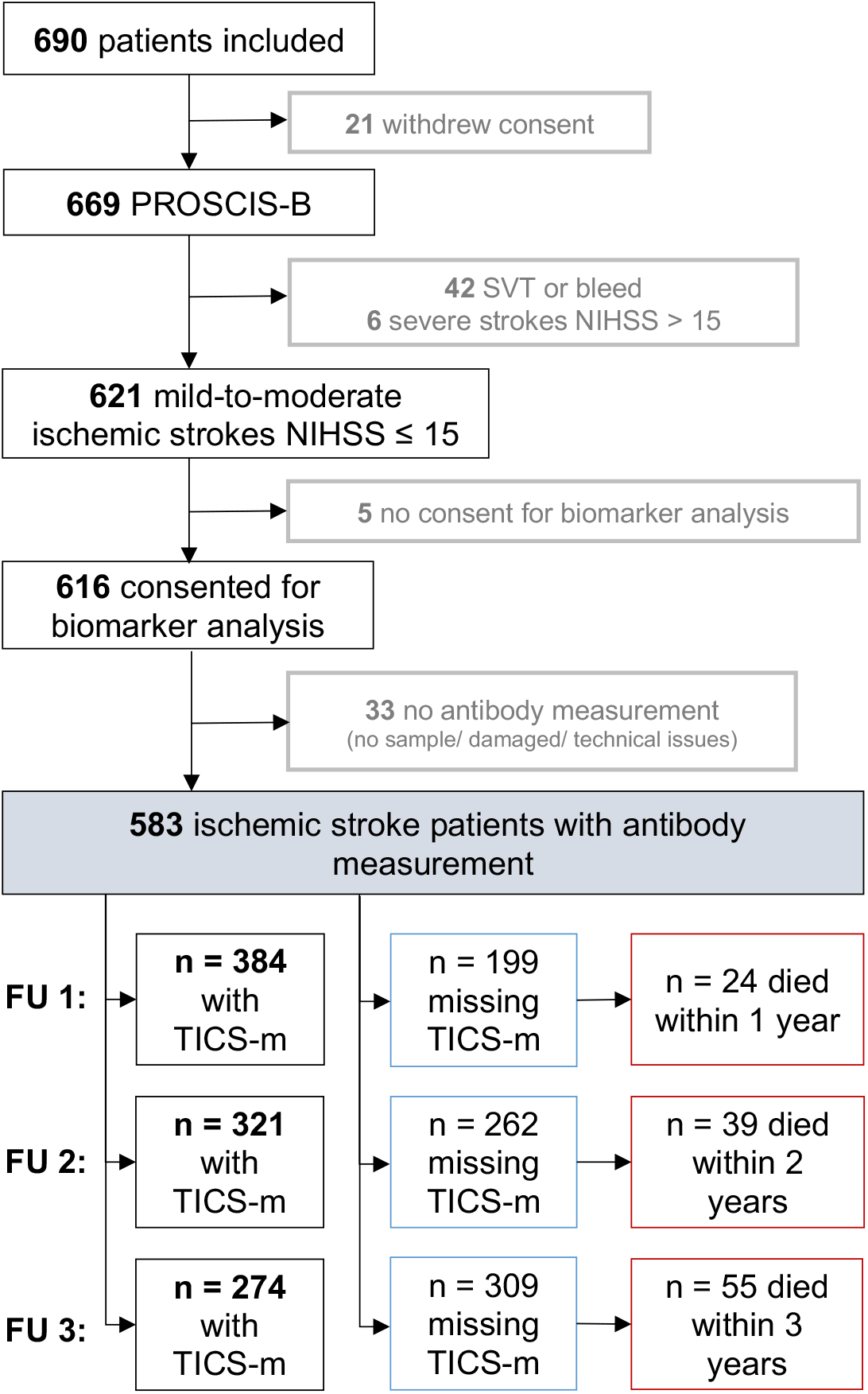
Flowchart of PROSCIS-B Inclusion and Exclusion and Overview on Follow-Up Data on Cognitive function.

PROSCIS-B patients had a mean age of 67 (Standard deviation [SD]=13), 39% were female, median baseline NIHSS was 2 (IQR=1 to 4) and baseline mRS was 2 (IQR=1 to 3). We measured NMDAR1-abs seropositivity in 76 patients (13%) in whom IgM NMDAR1-abs were present in serum of 49 patients (8%), followed by IgA (43 patients (7%)), and IgG isotype antibodies were detected in 2 patients (0.3%), only. Seventeen patients (3%) were presenting IgM and IgA antibodies, simultaneously. We did not observe major differences in baseline characteristics between NMDAR1-abs seropositive and seronegative patients and likewise not between subgroups as shown in Table 1. Cognitive function at baseline was assessed with the MMSE and was 28 points (IQR=26 to 30) on a median in the total cohort and in seronegative patients. In overall seropositive patients the median MMSE was 29 points (IQR=27 to 30) in seropositive patients with low titers the median MMSE was 29 points (IQR=27.5 to 30) and for those patients with high titers it was 27 points (IQR=24 to 29).

**Table 1.** Baseline Characteristics Table of PROSCIS – B Participants

|  | **PROSCIS – B** | **stratified by anti-NMDAR GluN1 antibody serostatus** |  |  |  |
| --- | --- | --- | --- | --- | --- |
|  | total | seronegative | seropositive | titer ≤ 1:100 | titer > 1:100 |
| **PROSCIS-B participants \* *n* (%)** | 621 (100) | 507 (82) | 76 (13) | 55 (9) | 21 (4) |
| anti-NMDAR GluN1 IgM antibodies *n* (%) | 49 (8) | – | 49 (8) | 34 (6) | 15 (3) |
| anti-NMDAR GluN1 IgA antibodies *n* (%) | 43 (7) | – | 43 (7) | 31 (5) | 12 (2) |
| anti-NMDAR GluN1 IgG antibodies *n* (%) | 2 (>0) | – | 2 (>0) | 2 (>0) | 0 |
| **age (years) mean (SD)** | 67 (13) | 67(13) | 66 (14) | 65 (14) | 71 (10) |
| **age (years) median (IQR)** | 69 (58 – 76) | 69 (59 – 76) | 67 (56 – 77) | 63 (51 – 77) | 69 (66 – 78) |
| **female sex *n* (%)** | 242 (39) | 204 (40) | 22 (29) | 17 (40) | 5 (24) |
| **systolic blood pressure (mmHg) mean (SD)** | 139 (22) | 139 (22) | 139 (24) | 139 (22) | 140 (28) |
| **diastolic blood pressure (mmHg) mean (SD)** | 77 (14) | 77 (15) | 78 (13) | 80 (12) | 73 (14) |
| **history of hypertension *n* (%)** | 406 (65) | 336 (66) | 46 (61) | 30 (55) | 16 (76) |
| **BMI (kg/m<sup>2</sup>) median (IQR)** | 27 (24 – 30) | 27 (24 – 29) | 28 (24 – 31) | 27 (24 – 30) | 30 (26 – 34) |
| **habitual alcohol consumption *n* (%)** | 217 (35) | 179 (36) | 23 (31) | 15 (27) | 8 (38) |
| **current smoker *n* (%)** | 171 (28) | 139 (28) | 22 (30) | 17 (31) | 5 (24) |
| **total cholestherine (mg/dl) mean (SD) ‡** | 198 (48) | 199 (48) | 198 (50) | 204 (51) | 180 (42) |
| **HDL (mg/dl) mean (SD) §** | 51 (16) | 52 (16) | 49 (17) | 50 (18) | 47 (13) |
| **LDL (mg/dl) mean (SD) §** | 122 (41) | 122 (41) | 124 (43) | 128 (43) | 112 (40) |
| **triglyceride (mg/dl) mean (SD) \|\|** | 139 (80) | 136 (80) | 152 (80) | 152 (80) | 152 (81) |
| **history of diabetes mellitus n (%)** | 137 (22) | 107 (21) | 21 (28) | 13 (24) | 8 (38) |
| **pAVK n (%)** | 42 (7) | 34 (7) | 6 (8) | 3 (6) | 3 (14) |
| **KHK n (%)** | 99 (16) | 80 (16) | 16 (21) | 10 (18) | 6 (29) |
| **history of atrial fibrillation n (%)** | 132 (21) | 106 (21) | 18 (24) | 11 (20) | 7 (33) |
| **estimated GFR (ml/min) mean (SD)** | 77 (21) | 77 (21) | 79 (22) | 83 (21) | 70 (22) |
| **NIHSS median (IQR)** | 2 (1 – 4) | 2 (1 – 4) | 3 (1 – 5) | 2 (1 – 5) | 3 (2 – 5) |
| **NIHSS: 0 – 4 n (%)** | 470 (76) | 386 (76) | 54 (71) | 40 (73) | 14 (67) |
| **NIHSS: 5 – 15 n (%)** | 151 (24) | 121 (24) | 22 (29) | 15 (27) | 7 (33) |
| **TOAST arterial atherosclerosis n (%)** | 167 (27) | 128 (25) | 25 (33) | 17 (31) | 8 (38) |
| **TOAST cardioembolic n (%)** | 145 (23) | 121 (24) | 18 (24) | 12 (22) | 6 (29) |
| **TOAST small vessel disease n (%)** | 96 (15) | 87 (17) | 6 (8) | 4 (7) | 2 (10) |
| **TOAST other n (%)** | 22 (4) | 15 (3) | 2 (3) | 2 (4) | 0 |
| **TOAST undetermined etiology *n (%)*** | 191 (31) | 156 (31) | 25 (33) | 20 (36) | 5 (24) |
| **mRS median (*IQR*)** | 2 (1 – 3) | 2 (1 – 3) | 2 (1 – 3) | 2 (1 – 3) | 2 (1 – 3) |
| **≤ 10 years of school education *n (%)*** | 421 (68) | 345 (72) | 51 (68) | 34 (63) | 17 (81) |
| **> 10 years of school education *n (%)*** | 171 (28) | 136 (28) | 24 (32) | 20 (37) | 4 (19) |
| **MMSE median (*IQR*)<sup>§</sup>** | 28 (26 – 30) | 28 (26 – 30) | 29 (27 – 30) | 29 (27.5 – 30) | 27 (24 – 29) |

SD, Standard deviation; IQR, inter quartile range between the 25<sup>th</sup> and 75<sup>th</sup> percentile; MI, myocardial infarction; PAD, peripheral artery disease; CHD, coronary heart disease; BMI, Body Mass Index; GFR, glomerular filtration rate calculated using the Chronic Kidney Disease Epidemiology Collaboration (CKD-EPI) formula; HDL, high density lipoprotein; LDL, low density lipoprotein; NIHSS, National Institutes of Health Stroke Scale; TOAST, stroke etiology according to Trial of Org 10172 in Acute Stroke Treatment; mRS, modified Rankin Scale; MMSE, Mini Mental State Examination \*38 participants were missing antibody measurements; Missing values were < 10% in all characteristics except for ‡‘total cholesterol’ missing: n = 57, §‘HDL’ and ‘LDL’ missing: n = 38 , ||‘Triglycerides’ missing: n = 49; Due to rounding values might not add to 100%.

Twenty-eight percent of PROSCIS-B patients with MMSE assessment (n=605) were cognitive impaired with test results of equal or less than 26 points. Among seronegative patients with MMSE and NMDAR1-abs measurement (n=497) this proportion was 29% and among seropositive patients with both assessments (n=72) 22% were cognitively impaired. Out of 52 patients with low titers and both assessments 15% of the patients were cognitively impaired at baseline and in patients with high titers this proportion was 40% (n=20).

TICS-m scores for all observations over time after stroke are visualized in Figure 2 and Figure 3, stratified by exposure groups. These graphs indicate no crude difference of cognitive performance over time between seropositive and seronegative patients. Patients with high titers do differ from seronegative patients and also from patients with low titers of NMDAR antibodies in cognitive outcome over years after stroke in the crude visualizations. We did not observe an effect of NMDAR1-abs seropositivity compared to seronegativity on TICS-m over time in the crude model (crude β=0.38 [95%CI: −1.00 to 1.76]) and in adjusted analyses (Model3 β=0.30 [95%CI: −1.14 to 1.73]). Time was associated with a positive effect on TICS-m scores of 0.71 points (95%CI: 0.45 to 0.96) in all patients. In patients with low titers of NMDAR1-abs, TICS-m over time was not different compared to seronegative patients in the crude (β=1.53 [95%CI: −0.06 to 3.11]) and full adjusted model (Model3 β=1.42 [95%CI: −0.23 to 3.08]). In patients with high NMDAR1-abs titers TICS-m over time was lower compared to seronegative patients in the crude model (β=-2.54 [95%CI: −4.99 to −0.08]) with a similar effect size in the full adjusted Model3 (β=-2.30 [95%CI: −4.82 to 0.21]). All effect estimates for NMDAR1-abs seropositivity exposure on TICS-m over time compared to NMDAR1-abs seronegativity are shown in Table 2. Effect estimates for all potential confounders including a quantification of the explained and residual variance within these models are presented in Supplemental Table I.

**Table 2.**
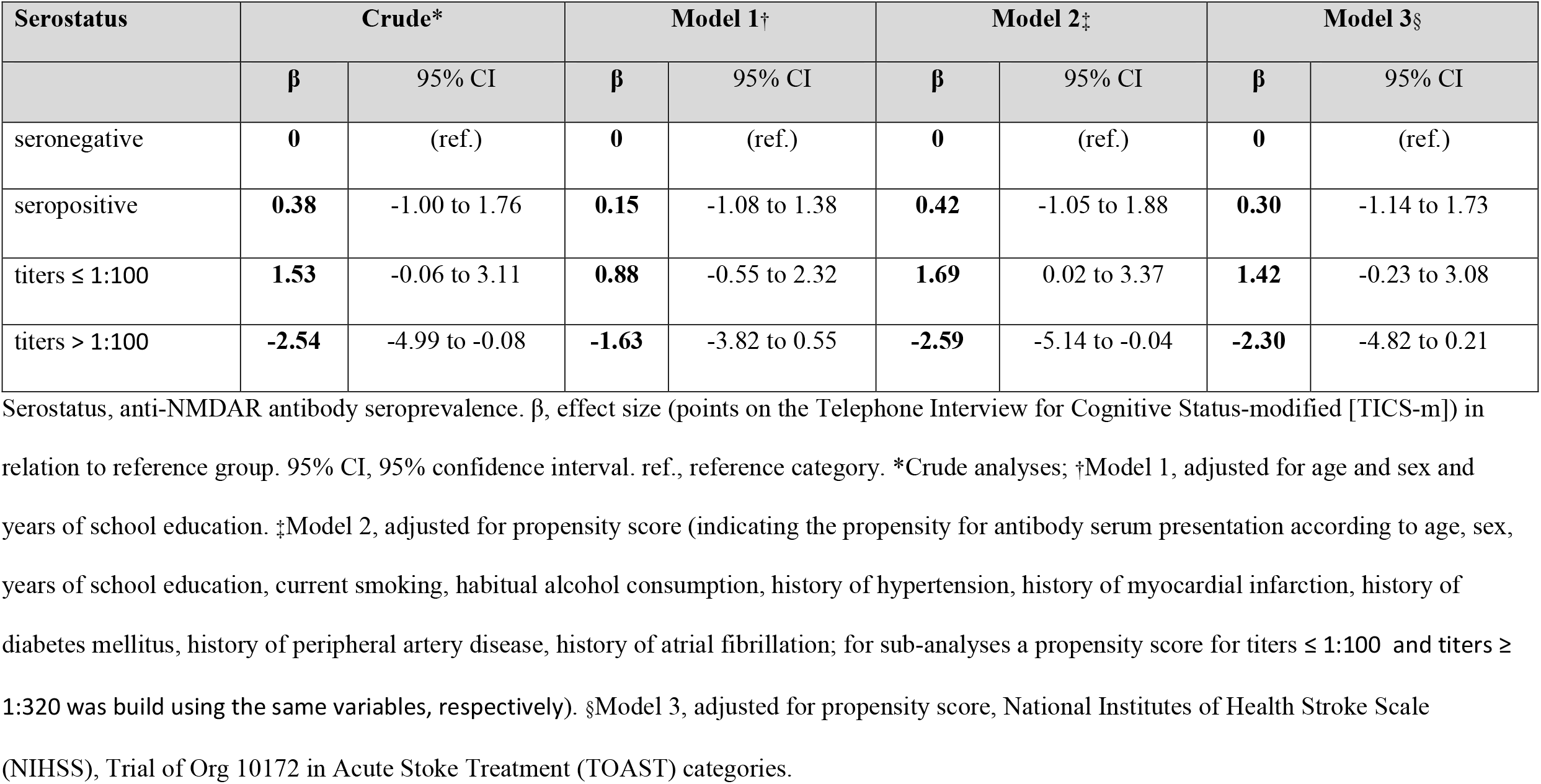
Anti-NMDA-Receptor Antibody Seropositivity and Cognitive Function Over Time after Stroke

**Figure 2:**
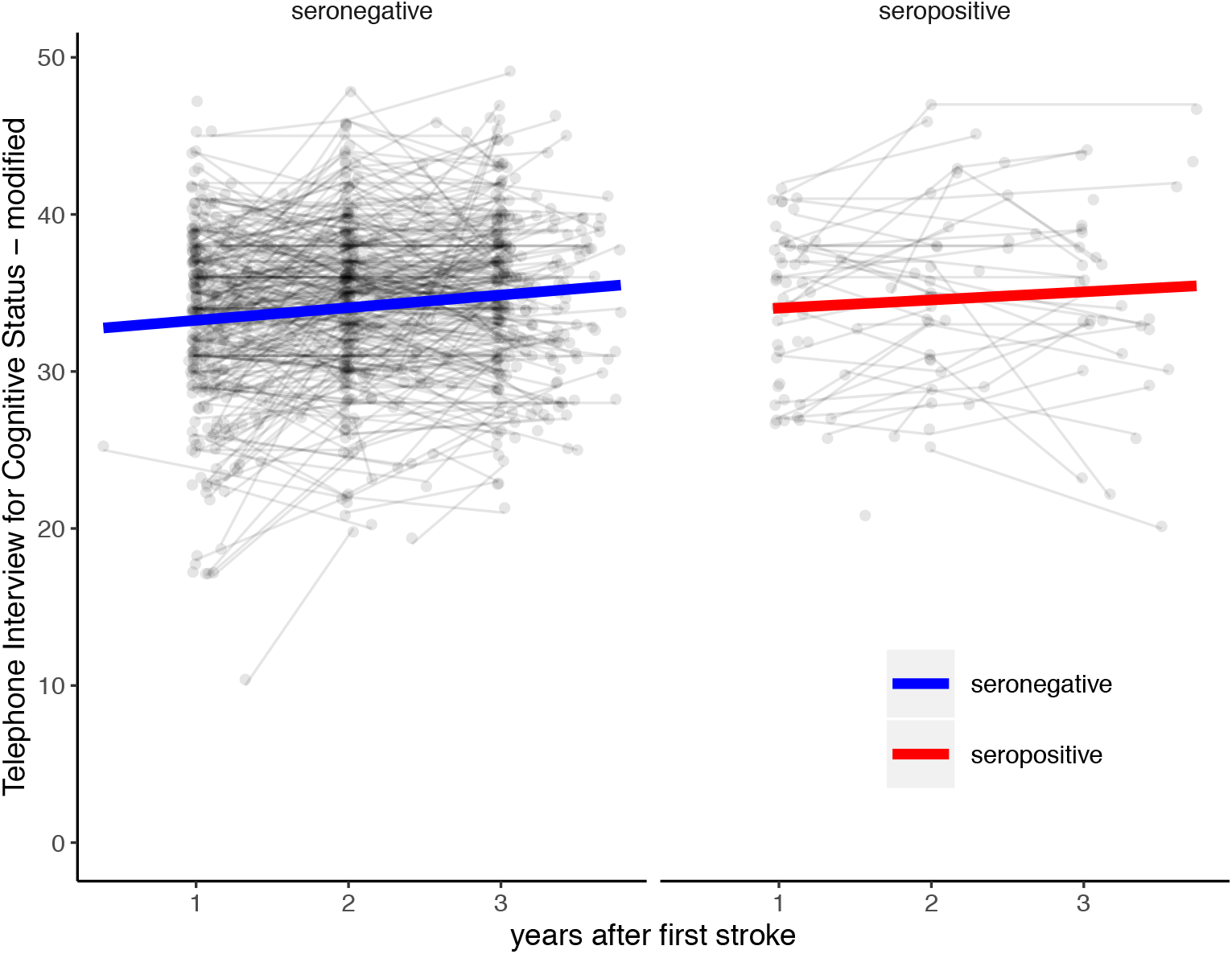
Anti−NMDA−Receptor GluN1 Antibody Seropositive and Seronegative Patients and Cognitive Function (TICS−m Scores) over Time after First Stroke.

**Figure 3:**
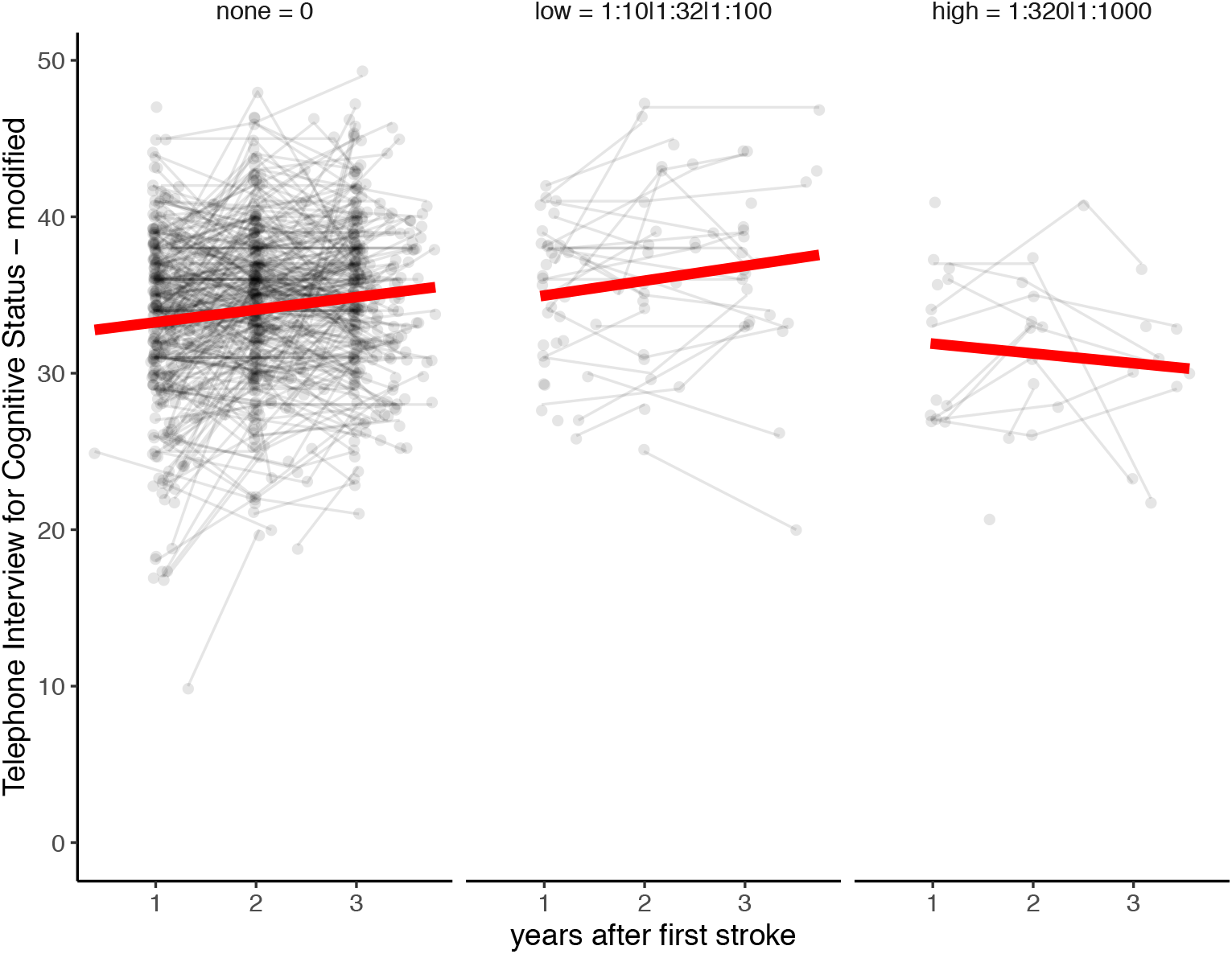
Cognitive Function (TICS−m Scores) over Time in Patients after First Stroke − stratified by Anti−NMDA−Receptor GluN1 Antibody Serum Titer Subgroups.

#### Results from sensitivity analyses

1) Comparing patients with at least one TICS-m follow-up assessment to patients with no follow-up TICS-m assessment at all, we observed that 14% of the patients with at least one TICS-m were seropositive compared to only 7% of patients with no TICS-m follow-up data. Some of the baseline characteristics indicated lower cardiovascular risk profile of patients with at least one TICS-m measurement (Current smokers: 36% compared to 26%; median MMSE: 29 [IQR:26 to 30] compared to 27 [IQR:25 to 29]). Full data are shown in Supplemental Table II. 2) After applying multiple imputation for 10 complete datasets, the pooled effect sizes appeared to be slightly attenuated but similar compared with the effect sizes of our main analysis. Results are presented in Supplemental Table III of the online only material. 3) Supplemental Table IV shows how many patients we excluded due to depression in each year given a best-case and worst-case scenario stratified by titer level subgroups. Effects for high titers of NMDAR1-abs on TICS-m over time were strongly attenuated after exclusion of observations from depressed patients in both, the best-case and worst-case scenario (Model3: BC β= −1.79; 95%CI: −4.64 to 1.06 / WC β= −1.12; 95%CI: −3.99 to 1.74). We provide full data in Supplemental Table V.

## Discussion

In our study of mild-to-moderately affected first-ever ischemic stroke patients, NMDAR1-abs seropositivity in the acute phase, primarily IgM and IgA, was not associated with cognitive function over three years after stroke. In subgroups, we also observed no differences between seronegative patients and patients with low titers. However, our data indicate decreased cognitive function over time after ischemic stroke in patients with high titers of NMDAR1-abs compared to seronegative patients, which is independent from several other risk factors. NMDAR1-abs serum prevalence has been observed in about ∼10% of the healthy population, and in those with various diseases, including stroke.(9,10,20) Similar to our study results, mainly IgM and IgA antibodies were observed.(10) The swift blood sampling in our study and the similar proportion of seropositive patients in healthy individuals suggests that antibodies were preexistent before the stroke and thus not a consequence of the acute event.(20) This is further supported by findings from the study of Zerche et al. who measured NMDAR1-abs in a more standardized time-window swiftly after the stroke (<36h) and measured a similar serum prevalence, using the same determination technique like we used for our study.(7) The high serum prevalence of IgM and IgA NMDAR1-abs in healthy elderly individuals questions a pathological significance of these isotypes on their own. Cumulative evidence from disease cohorts other than stroke patients link serum presentation particularly with cognitive outcomes: for example was NMDAR1-abs IgA and IgM seropositivity associated with cognitive impairment in melanoma patients, and others found serum IgA to be associated with different types of slowly progressive cognitive impairment.(3–5) In stroke, however, NMDAR1-abs seropositivity was foremost linked with beneficial effects on infarct volumes, which was explained by presumed effects of NMDAR1-abs on NMDA-receptor mediated excitotoxicity.(7,8,21) In line with other studies mentioned earlier,(3–5) in our study of first-ever stroke patients high titers were associated with decreased cognitive function, challenging previous hypotheses of beneficial effects of these antibodies in stroke pathology.(7,8) This current observation further coincides with our observation regarding NMDAR1-abs and other outcomes in the PROSCIS study, i.e. increased vascular risk for overall seropositive patients and unfavorable functional outcome in patients with high titers, after stroke respectively.(6) Considering that NMDARs are highly expressed in the central nervous system and that the blood brain barrier disruption in stroke allows serum NMDAR1-abs to access vulnerable neuronal tissue, it appears reasonable to assume antigen binding of NMDAR1-abs to their targeted receptor and that they subsequently exert functional effects. If binding results in NMDAR downregulation, as proposed previously,(10,22) this could hamper functional recovery and lead to impaired tissue reorganization long-term after stroke.(23) This is because NMDARs are crucial for mechanisms of synaptic plasticity and normal neuronal functioning.(11,21) Although different groups reported functional effects of IgA and IgM on NMDARs in terms of receptor internalization, the question whether these effects are sufficient to have clinical implications remains a controversy.(4,9,10) NMDAR1-abs may also be markers of an underlying immunology which associates with vascular risk, functional and neuropsychiatric impairment and may therefore be predictive biomarkers for outcomes after stroke, irrespective of an underlying causal relationship. Either way our effect sizes combined with the precision of estimation is not indicative for the quality of an underlying relationship. For more clarification, our study would have benefitted from a deep immune-characterization of study subjects. We could not observe any associations of NMDAR1-abs serum presentation with low titers and cognitive outcome. If we take into account that only high titers associate with unfavorable functional outcome after stroke, a higher cut-off for seropositivity should be considered.(6)

We consider the effects observed for high NMDAR1-abs titers as clinically significant, supported by its effect size, representing half of a SD of overall TICS-m result (SD of TICS-m at the first year after stroke = 5.4 points).(24) If we further take a look at the crude effect for stroke severity (NIHSS 5-15 vs. 0-4) on TICS-m, which was −2.12 points (95%CI: −3.19 to −1.05) in the PROSCIS-B study and thus essentially the same effect than the one we observed for NMDAR1-abs seropositivity with high titers on TICS-m, the importance may become clearer. Overall, our Model2 may be the most reliable estimate because we comprehensively adjusted for potential confounders while incorporating only a single parameter, i.e. the propensity score, which prevented over-parametrization of the model. In a sensitivity analysis, we excluded TICS-m observations of depressed patients and effects observed for seropositivity with high titers were extremely attenuated. This is explained by the exclusion of a high proportion of seropositive participants with high titers (see Supplemental Table IV) and raises the question whether those excluded patients suffered both, i.e. cognitive impairment and depression, or whether pseudodementia due to depression was present in these patients and cognitive decline played a minor role. Our cognitive outcome scores were increasing over time indicated by a precise positive effect size for time in our models (see Supplemental Table I), which we interpret as a learning effect for TICS-m due to repeated measures.(25) Otherwise, our models seem to accurately pick up on the effects, as demonstrated by single estimates for our prechosen explanatory variables: for example, higher school education (>10 years) caused an increase of 3.57 TICS-m points compared to lower school education (<= 10 years) (Supplemental Table I), which supports that despite this learning effect our study remains representative.

### Limitations

Our study results may be biased towards the null due to a learning effect in the TICS-m instrument. This consideration was previously reported for TICS-m in other studies.(25) Furthermore, we were not able to conduct a detailed neuropsychological testing, which could discriminate even minor disabilities in patients with only mild-to-moderate stroke events. To reduce internal heterogeneity we excluded those six patients with a baseline NIHSS of ≥16. However, differences in cognitive function may be more pronounced in severely affected stroke patients. Furthermore, our study results cannot be generalized to patients with severe stroke events. Selection bias may be present, as we recorded a total of ∼ 40% of the observations were missing TICS-m assessments. We addressed this short-coming in two sensitivity analyses, one of which was a multiple imputation procedure for missing values. However, as we cannot exclude that patients were missing an assessments not at random bias may still be present or even worsened.(26) Our results need further validation, especially because our effect estimate did not appear robust in different sensitivity analyses. Besides these limitations, this study followed an a priori analysis plan with prespecified confounder adjustment and we benefit from highly standardized state-of-the-art antibody measurements.(27) We revisited our study results with several sensitivity analyses, which elucidated that our study results demand further validation.

To conclude, NMDAR1-abs overall seropositivity and NMDAR1-abs was not associated with cognitive function over time after first-ever ischemic mild-to-moderate stroke. Our data point towards a relevance of high serum titers (>1:100) of IgA and IgM NMDAR1-abs as biomarkers for unfavorable cognitive outcome after stroke. The causal nature of the relationship between high titers of NMDAR1-abs and worse cognitive functioning should be subject of future experimental and clinical investigations.

## Acknowledgements

The authors want to thank the EUROIMMUN laboratory for NMDAR1-abs measurements and Jane Thümmler for data management. PSS is a MD candidate at the Charité – Universitätsmedizin Berlin, and this work is submitted in partial fulfillment of her thesis.

## References

1. Mijajlovi MD, Pavlovi A, Brainin M, et al. Post-stroke dementia – a comprehensive review. BMC Medicine. 2017;1–12. doi:10.1186/s12916-017-0779-7

2. Pendlebury ST, Rothwell PM. Incidence and prevalence of dementia associated with transient ischaemic attack and stroke: analysis of the population-based Oxford Vascular Study. Lancet Neurol. 2019;18(3):248–58. doi:https://doi.org/10.1016/S1474-44221830442-3

3. Doss S, Wandinger KP, Hyman BT et al. High prevalence of NMDA receptor IgA/IgM antibodies in different dementia types. Ann Clin Transl Neurol. 2014;1(10):822–32. doi:10.1002/acn3.120.

4. Prüss H, Höltje M, Maier N et al. IgA NMDA receptor antibodies are markers of synaptic immunity in slow cognitive impairment. Neurology. 2012;78(22):1743–53. doi: 10.1212/WNL.0b013e318258300d

5. Bartels F, Strönisch T, Farmer K et al. Neuronal autoantibodies associated with cognitive impairment in melanoma patients. Ann Oncol Off J Eur Soc Med Oncol. 2019;30(5):823–9. doi:10.1093/annonc/mdz083.

6. Sperber PS, Siegerink B, Huo S et al. Serum Anti-NMDA (N-Methyl-D-Aspartate)-Receptor Antibodies and Long-Term Clinical Outcome After Stroke (PROSCIS-B). Stroke. 2019;1–7. doi:10.1161/STROKEAHA.119.026100.

7. Zerche M, Weissenborn K, Ott C et al. Preexisting Serum Autoantibodies Against the NMDAR Subunit NR1 Modulate Evolution of Lesion Size in Acute Ischemic Stroke. Stroke. 2015;46(5):1180–6. doi:10.1161/STROKEAHA.114.008323.

8. During MJ, Symes CW, lawlor PA, et al. An Oral Vaccine Against NMDAR1 with Efficacy in Experimental Stroke and Epilepsie. Science. 2000;2716(February). doi:10.1126/science.287.5457.1453

9. Hara M, Martinez-Hernandez E, Ariño H, Armangué T et al. Clinical and pathogenic significance of IgG, IgA, and IgM antibodies against the NMDA receptor. Neurology. 2018;90(16):E1386–94. doi:10.1212/WNL.0000000000005329.

10. Castillo-Gómez E, Oliveira B, Tapken D et al. All naturally occurring autoantibodies against the NMDA receptor subunit NR1 have pathogenic potential irrespective of epitope and immunoglobulin class. Mol Psychiatry. 2017;22(12):1776–84. doi:10.1038/mp.2016.125.

11. Paoletti P, Bellone C, Zhou Q. NMDA receptor subunit diversity: impact on receptor properties, synaptic plasticity and disease. Nat Rev Neurosci. 2013;14(6):383–400 doi: https://doi.org/10.1038/nrn3504.

12. Hatano S. Experience from a multicentre stroke register: a preliminary report. Bull World Health Organ. 1976;54(5):541–53.

13. Liman TG, Zietemann V, Wiedmann S et al. Prediction of vascular risk after stroke - protocol and pilot data of the prospective Cohort with incident stroke (PROSCIS). Int J Stroke. 2013;8(6):484–90. doi: 10.1111/j.1747-4949.2012.00871.x.

14. Malsch C, Liman T, Wiedmann S, et al. Outcome after stroke attributable to baseline factors — The PROSpective Cohort with Incident Stroke (PROSCIS). PLoS One. 2018;13(9):1–14. doi: 10.1371/journal.pone.0204285.

15. Ramberger M, Peschl P, Schanda K et al. Comparison of diagnostic accuracy of microscopy and flow cytometry in evaluating N-methyl-D-aspartate receptor antibodies in serum using a live cell-based assay. PLoS One. 2015;10(3):1–18. doi: 10.1371/journal.pone.0122037.

16. Dalmau J, Tüzün E, Wu HY et al. Paraneoplastic anti-N-methyl-D-aspartate receptor encephalitis associated with ovarian teratoma. Ann Neurol. 2007;61(1):25–36. doi:10.1002/ana.21050

17. Pendleburry ST, Markwick A, de Jager CA, et al. Differences in Cognitive Profile between TIA, Stroke and Elderly Memory Research Subjects?: A Comparison of the MMSE and MoCA. Cerebrovasc Dis. 2012;48–54. doi:10.1159/000338905.

18. Knopman DS, Roberts RO, Geda YE et al. Validation of the telephone interview for cognitive status-modified in subjects with normal cognition, mild cognitive impairment, or dementia. Neuroepidemiology. 2010;34(1):34–42. doi:10.1016/j.archger.2010.04.008.

19. Clark MH. Propensity Scoring. International Encyclopedia of Social & Behavioral Sciences. 2015.140–146p. doi:10.1016/B978-0-08-097086-8.10557-4

20. Dahm L, Ott C, Steiner J et al. Seroprevalence of autoantibodies against brain antigens in health and disease. Ann Neurol. 2014;76(1):82–94. doi:10.1002/ana.24189.

21. Hardingham GE, Bading H. Synaptic versus extrasynaptic NMDA receptor signalling: implications for neurodegenerative disorders. Nat Rev Neurosci. 2010;11(10):682–96. doi:10.1038/nrn2911

22. Kreye J, Wenke NK, Chayka M et al. Human cerebrospinal fluid monoclonal N - methyl-D-aspartate receptor autoantibodies are sufficient for encephalitis pathogenesis. Brain. 2016;2641–52. doi:10.1093/aww213

23. Ikonomidou C, Turski L. Personal view Why did NMDA receptor antagonists fail clinical trials for stroke and traumatic brain injury?? Lancet Neurol. 2002;1(6):383–6. doi:10.1016/s1474-44220200164-3

24. Leppink J, O’Sullivan P, Winston K. Effect size – large, medium, and small. Perspect Med Educ. 2016;5(6):347–9. doi:10.1007/s40037-016-0308-y

25. Katan M, Wright CB, Gardener H, Dong C, Marquez C, DeRosa JT, et al. Infectious burden and cognitive performance: The northern Manhattan study. Stroke. 2013;45. doi:10.1212/WNL.0b013e3182896e79.

26. Sterne JAC, White IR, Carlin JB, et al. Multiple imputation for missing data in epidemiological and clinical research: Potential and pitfalls. BMJ. 2009;339(7713):157–60. doi:10.1136/bmj.b2393.

27. Waters P, Reindl M, Saiz A, et al. Multicentre comparison of a diagnostic assay: Aquaporin-4 antibodies in neuromyelitis optica. J Neurol Neurosurg Psychiatry. 2016;87(9):1005–15. doi:10.1136/jnnp-2015-312601.

